# COVID-19 Pandemic and Prevalence of Self-care Practices among the Future Physicians: A Bangladesh Study

**DOI:** 10.1101/2021.05.11.21257027

**Authors:** Fatema Johora, Asma Akter Abbasy, Fatiha Tasmin Jeenia, Mithun Chandro Bhowmik, Mohsena Aktar, Nargis Akhter Choudhury, Priyanka Moitra, Jannatul Ferdoush

## Abstract

**Background:** Since December 2019, the novel coronavirus, SARS-CoV-2, has garnered global attention due to its rapid transmission, which has infected more than twenty nine million people worldwide. World is facing enormous stress and anxiety as there is no effective medicine or vaccine to treat or prevent COVID-19 till date. Experts are recommending self-care like social distancing, respiratory etiquette, hand washing, using face mask to prevent corona virus infection.

**Materials and methods:** This descriptive cross-sectional study was designed to assess the prevalence of self-care practice among the undergraduate medical students (4^th^ year) of 14 medical colleges of Bangladesh during COVID-19 pandemic. A structured questionnaire survey linked in the google form was used as study instrument and was distributed among study population through email, messenger, whatsapp and other social media during the month of October 2020. Total 916 students were participated in the study.

**Results:** 79.8% of students reported self-care practice in study period. 44.98% of students went outside once in a week. 90.5%, 70.96% and 52.62% of respondents always used face mask, followed 20 seconds hand washing principle and maintained social distancing. Face masks (97.8%), sanitizers (76.7%) and gloves (71.9%) are most common items purchased as protective mesures. Most of the students (76.9%) follow their hobbies as a coping strategy to overcome phychological stress, while 6% of students took professional help.

**Conclusion:** Suboptimal practice of self-care was found among the undergraduate medical students of Bangladesh.

## INTRODUCTION

The outbreak of corona virus disease (COVID-19) becomes a global crisis. In December 2019, few cases of atypical pneumonia were reported in Wuhan, China and later identified as novel coronavirus infection caused by Severe acute Respiratory Syndrome Coronavirus 2 (SARS-CoV-2).^[1, 2]^ On January 30, 2020 WHO declared it’s as a global health emergency and later as pandemic on March 11, 2020. By 17 September 2020, total 216 coutries, areas or territories are affected with case, 29679284 are already infected and 936521 died because of COVID-19 pandemic.^[3, 4]^ In Bangladesh, first case of COVID-19 was detected on 8 March 2020 and rapidly spread out throughout the country.^[5]^ The COVID-19 causes a plethora of clinical manifestations, and the severity and outcomes may vary, depending on the underlying co-morbidities like diabetes, heart diseases, hypertension, COPD, bronchial asthma as well as age, sex, and geographic locations of the patients.^[6]^ The rising rates of cases are putting healthcare system in jeopardy.^[7, 8]^ Several therapeutic agents have been used for the treatment of COVID-19. However, there is no effective antiviral drug or vaccine to combat COVID-19 pandemic.^[9, 10]^ As it’s a highly contagious disease, non-pharmaceutical interventions (NPIs) remain the primary strategy to control COVID-19. The NPIs include public health control measures such as quarantine, travel restrictions, isolation, social distancing, and infection prevention and control to reduce transmission of the disease. Several impact assessment studies have shown that public health interventions are associated with the reduced transmission of COVID-19.^[11, 12, 13]^ Prevention of infection control is one of the pivotal measures in the absence of an effective vaccine or treatment.^[14]^ World health organization has been repeatedly emphasizing this strategy through self-care practice.^[15]^

Self-care is the ability of individuals, families and communities to promote health, prevent disease, maintain health and to cope with illness and disability with or without the support of a healthcare provider. It encompasses several issues including hygiene, nutrition, lifestyle, environmental, socio-economic factors and self medication. Promotion of self care is a means to empower individuals, families and communities for informed health decision-making. It has the potential of improving the efficiency of health systems and contributing towards health equity.^[16, 17]^ In a humanitarian or crisis situation like Covid-19 pandemic, self-care could play an important role to improve health-related outcome.^[18]^ Practicing of responsible self-care is crucial in decreasing the burden on healthcare systems through self-management of mild symptoms as well as implementation of preventive measures particularly during this time of crisis.^[19]^ Physical distancing, good respiratory hygiene and hand washing are important examples of self-care actions those has been promoted by WHO every day to protect individual from COVID-19, and there are many other areas in which self care can contribute to maintain physical and mental wellbeing during the coronavirus disease pandemic.^[17]^ Education of individual person is impotant to get fruitful effects of self-care practice as irresponsible practice can bring numerous potential hazards.^[19, 20]^ From the beginning of COVID-19 pandemic, world has been observing a pile of myths in mass media and social media.^[21, 22]^ Practice of self-care is influenced by several factors like education, gender, socioeconomic status and availability of medicine.^[23]^ But there is no evidence regarding self-care practice among the future physicians.

Hence, the present study will be carried out with the attempt to find out the prevalence of self-care practice among the undergraduate medical students of Bangladesh during Covid-19 pandemic. Finding of this research will be helpful to develop an educational intervention program on self-care practice will be integrated in order to improve rational use of medicine among the future physicians.

## Material & Methods

The objective of the study was to describe pattern of self-care practice among the undergratuate medical students of Bangladesh during COVID-19 pandemic.

### Study Design and Population

A descriptive cross-sectional study was designed to meet the study objective. The study population comprised of 4th year students of fourteen medical colleges of Bangladesh including government (Armed Forces Medical College, Cumilla Medical College, Colonel Malek Medical College, Manikganj and Rangpur Medical College) and non-government medical colleges (Army Medical College Bogura, Army Medical College Chattogram, BGC Trust Medical College, Brahmanbaria Medical College, Chattogram International Medical College, Jalalabad Ragib-Rabeya Medical College, Sylhet, Jashore Medical College, Khawja Yunus Ali Medical College, Sirajganj, Medical College for Women, Dhaka and US-Bangla Medical College, Narayanganj) in October 2020. Total 916 students were participated in the study.

### Study Instrument

A structured questionnaire was used for data collection and questionnaire was validated before survey.

### Procedure

Ethical approval was taken from the Institutional Review Board (IRB) of BGC Trust Medical College, Chittagong. Permission was taken from college authorities and informed consent was taken from the participants of the Structured Questionnaire Survey. Researchers explained the nature and purpose of the survey to the students during a virtual class. This self-administered questionnaire was linked in google form and was distributed among study population through email, messenger, whatsapp and other social media who gave consent. To assure the quality, students filled and submitted the questionnaire quickly during end of class. Later, this web-based questionnaire was sent to students who were absent in the class through email. A reminder mail or message was given on 7^th^ day and 15^th^ day of the primary one. The response generated by the students was received through google drive, and it did not accept double response from same participant. To maintain confidentiality, responses were anyoymus.

### Statistical analysis

Data was compiled, presented and and analyzed using Microsoft Excel 2007, and was expressed as percentage.

## Results

Nine hundreds and sixteen respondents were covered during the study period, of which 326 (35.59%) were males and 590 (64.41%) were females. Total 305 (33.23%) students responded that anyone in family including himself was diagnosed as COVID-19 and among them 242 (26.42%) were RT-PCR positive **(Table I)**. 731 (79.8%) students reported self-care practice in study period. 412 (44.98%) students went outside once in a week. 829 (90.5%) respondents always used facemask while go outside. 650 (70.96%) students always followed 20 sec hand washing principle, while 482 (52.62%) students always maintained social distancing **(Table II)**.

**Table I:** Demographic data.

| Variables |  | Frequency | Percentage |
| --- | --- | --- | --- |
| Gender | Male | 326 | 35.59% |
|  | Female | 590 | 64.41% |
| Diagnosed case of COVID-19 in family |  | 305 | 33.23% |
| RT-PCR positive in family |  | 242 | 26.42% |

**Table II:**
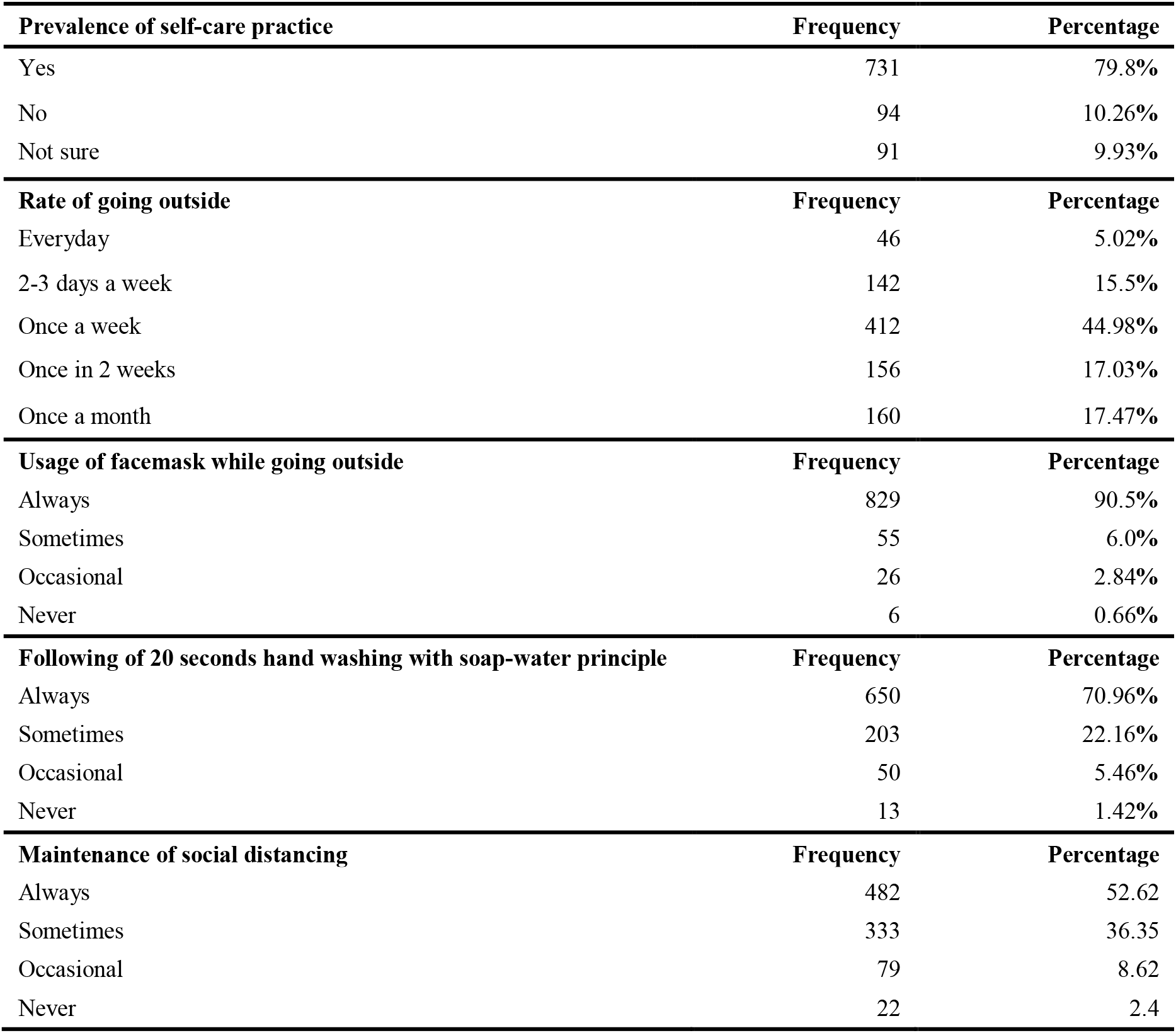
Responses of medical students about different aspects of self-care practice in last 06 months.

| Prevalence of self-care practice | Frequency | Percentage |
| --- | --- | --- |
| Yes | 731 | 79.8% |
| No | 94 | 10.26% |
| Not sure | 91 | 9.93% |
| **Rate of going outside** | **Frequency** | **Percentage** |
| Everyday | 46 | 5.02% |
| 2-3 days a week | 142 | 15.5% |
| Once a week | 412 | 44.98% |
| Once in 2 weeks | 156 | 17.03% |
| Once a month | 160 | 17.47% |
| **Usage of facemask while going outside** | **Frequency** | **Percentage** |
| Always | 829 | 90.5% |
| Sometimes | 55 | 6.0% |
| Occasional | 26 | 2.84% |
| Never | 6 | 0.66% |
| **Following of 20 seconds hand washing with soap-water principle** | **Frequency** | **Percentage** |
| Always | 650 | 70.96% |
| Sometimes | 203 | 22.16% |
| Occasional | 50 | 5.46% |
| Never | 13 | 1.42% |
| **Maintenance of social distancing** | **Frequency** | **Percentage** |
| Always | 482 | 52.62 |
| Sometimes | 333 | 36.35 |
| Occasional | 79 | 8.62 |
| Never | 22 | 2.4 |

**Figure 1** showed that facemasks (97.8%) and sterilizers, sanitizers (76.7%) are most common items purchased as protective mesures, followed by gloves (71.9%).

**Figure 1:**
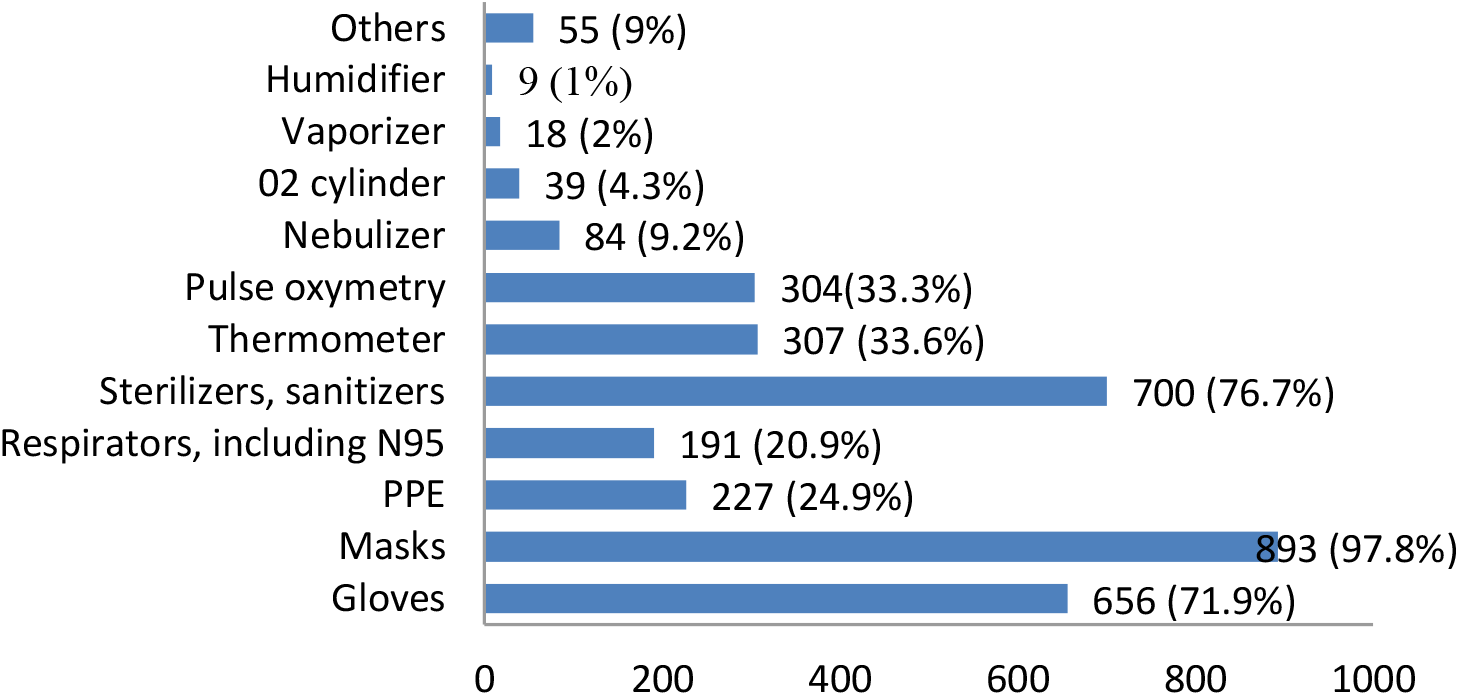
Items purchased as protective measures.

**Figure 2** showed that most of the students (76.9%) follow their hobbies to keep up mental health status, only 55 (6%) students took professional help.

**Figure 2:**
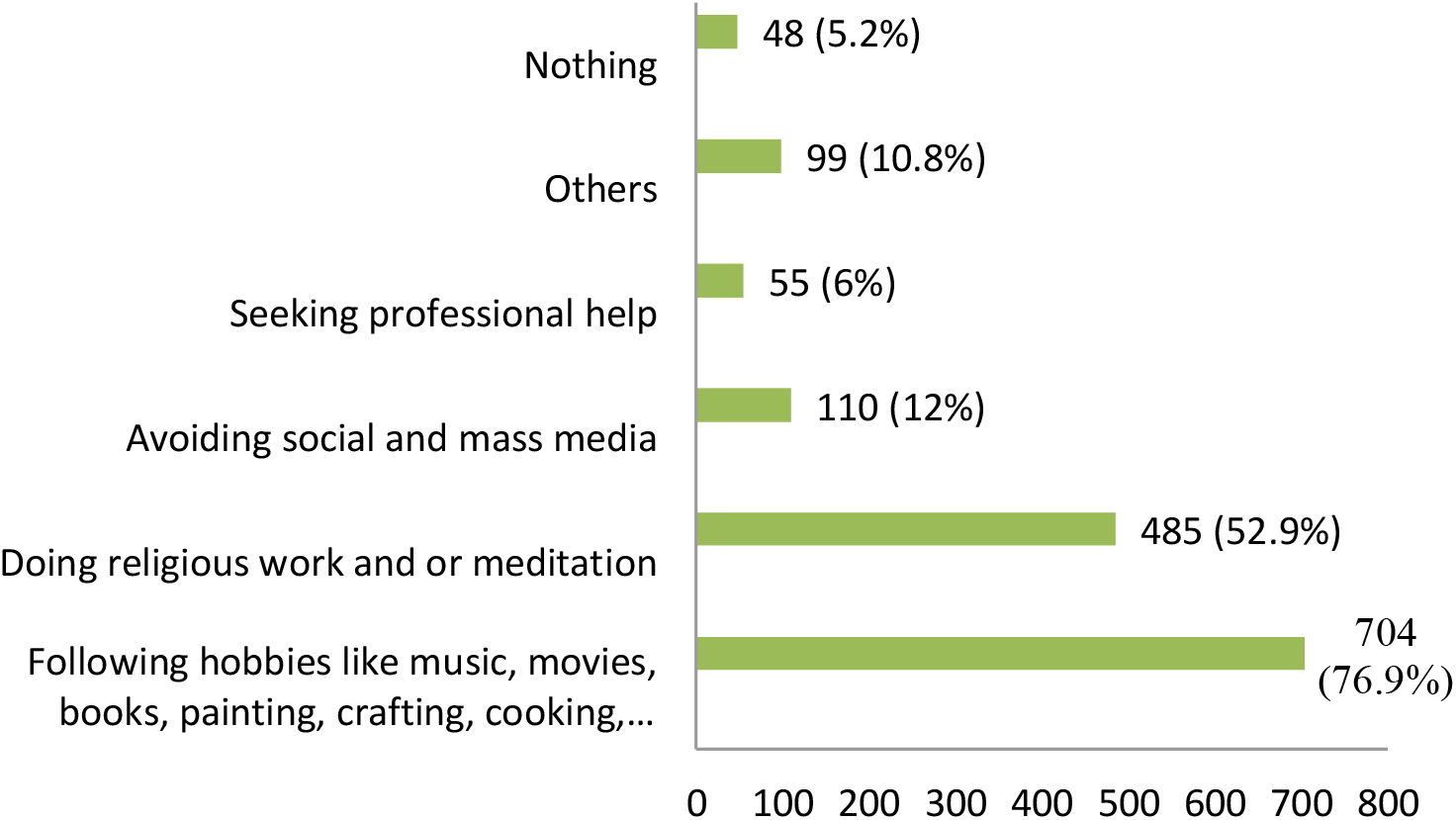
Measures taken for psychological and emotional well-being.

226 (25%) students participated online course on coronavirus deveoped by Diretorate General Of Health Service (DGHS) **(Figure 3)**.

**Figure 3:**
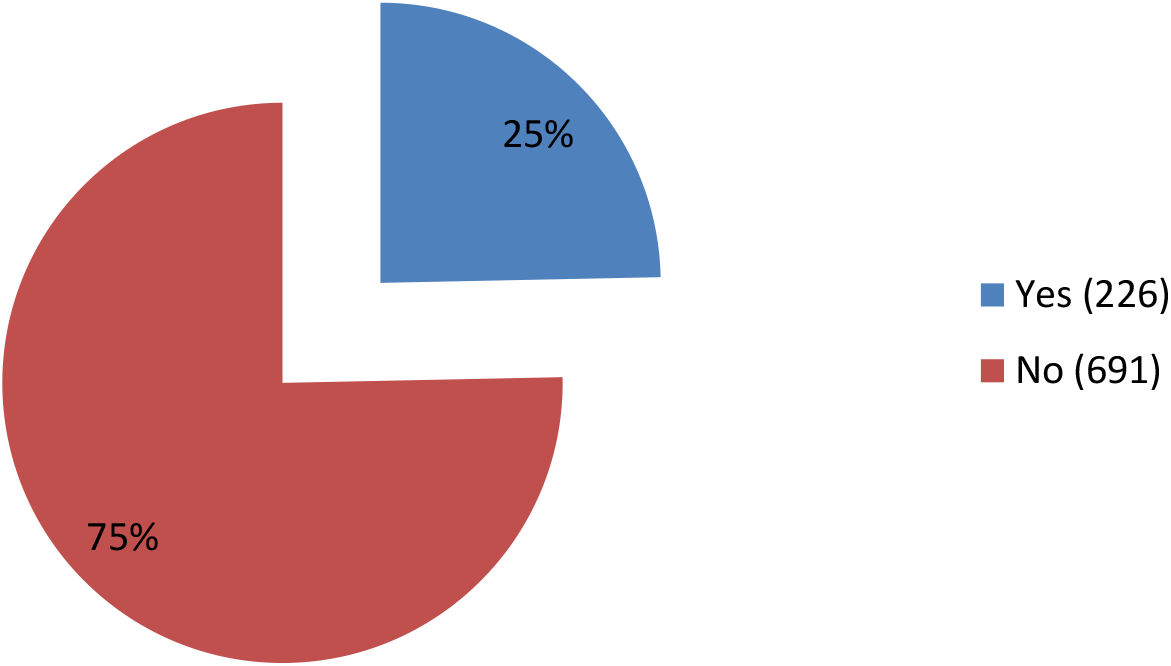
Participating online course on Coronavirus disease.

## Discussion

For last several months, world has been observing Covid-19 pandemic with enormous stress and anxiety. Still now, there is no effective medicine or vaccine to combat COVID-19. Lots of misonception are going on about preventive measures to control this higly contagious microorganisms. Current study was conducted in this context to explore self-care practices among undergraduate medical students of Bangladesh.

Social distancing, good respiratory hygiene and hand washing have been recommended by experts as self-care practice to prevent corona virus infection. 79.8% students responded about practicing self-care pratice in last 6 months but it was expected that everyone could practice self-care as they are previlieged with knowledge of medicine as well as information technology, may be a certain proprtion of students were not familiar with the term ‘self-care’. Most of the students followed ‘stay home principle’ to avoid chance of infection and this finding was similar to another study conducted among medical students.^[24]^

Although there were some controversies at the beginning, current best evidence suggests that facemask is beneficial for protection of both healthcare professionals and the public as wearing of mask reduces the transmissibility per contact by reducing transmission of infected droplets along with personal protection for the mask wearer. It is considered as most effective way to control spread of virus when compliance is high among larger population. ^[25, 26, 27]^ But use of masks is an evolving and cultural phenomenon, varies widely from country to country. ^[28, 29]^ In our study, most of the participants reported using of face mask always while going outside that was similar to two recent studies conducted in Bangladesh, ^[30, 31]^ poor practices were observed in studies conducted on pharmacy and medical students, ^[24, 32]^ probably cultural variation was the reason as those studies were conducted in Egypt and Jordan.^[28, 29]^. Hand hygiene is a key to prevent the spread of SARS-Cov-2. 20 seconds hand washing with soap water is considered as one of the most cost-effective way to mitigate COVID-19.^[33]^ Social distancing measures are other important tools to reduce the morbidity and mortality of COVID-19 to some degrees, and it seems to be crucial to control the pandemic, especially in the absence of definitive treatments and vaccines.^[34, 35]^ Suboptimal practices was found among undergraduate medical students of Bangladesh and this was contrary to similar studies ^[24, 31, 32]^

The extended practice of self-care among individuals has contribued to better health and more sustainable healthcare systems, and COVID-19 pandemic has prompted ths trend. Throughout the pandemic, many self-care product have been recommended to help threat the symptoms of coronovirus. ^[15, 18]^ In current study, masks, gloves, sanitizers, pulse oxymetery, thermometers were bought as protective measures, and these buying behavior reflects the extended practice of self-care. From the very beginning of pandemic, people started to buy daily necessities as well as medicine and equipments^[36, 37]^, and study showed relationship of higher level of education with increased buying behavior.^[29]^

COVID-19 pademic has caused major disruption of education systems worldwide including abrupt shut down of medical schools. Sudden changes in curricular delivery, loss of peer interaction, social isolation, uncertainity, financial stress all are negatively affecting mental well being of medical students.^[38]^ Most of the students resondended following their hobbies, religious works, meditation, where few of them seek professional help to cope up the stress. And these kind of activities were used as coping strategies among medical students. ^[38]^ Diretorate General of Health Services (DGHS) introduced an online course to orient and provide useful information to healthcare professionals as well as general population on the COVID-19 with the aim to provide optimum services to COVID-19 patients while ensuring one’s own protection and safety.^[39]^ Poor participation of medical students in this course might be because of their less famlilarity and acceptance with distance learning process.^[40]^

Undergraduate medical students are the ‘future physicians’, so it was expected that they would adhere completely with self-care practice to prevent transmission of coronavirus but suboptimal practice of self-care was found in this study.

## Conclusion

Self-care practices as usage of face masks, hand hygiene, social distancing, taking care of individual’s mental health are crucial measures to mitigate mortality and morbidity of COVID-19. In current study, suboptimal practices of these preventive measures were found among the undergratuate medical students of Bangladesh. Educational intervention on self-care practice might be helpful in improving current practices.

## Data Availability

All Data are available on request

